# Limited incremental value of three complete-blood-count inflammatory indices for cardiovascular mortality: a development and external validation study in 30,533 US adults

**DOI:** 10.64898/2026.09.30.26364394

**Authors:** Pengfei Guo, Wenlong Yan, Haoyu Hu, Yuan An, Yalin Liu, Sumin Yang

## Abstract

Routine blood tests can be used to calculate several inflammation-related ratios, including the neutrophil-to-lymphocyte ratio (NLR), systemic immune-inflammation index (SII), and platelet-to-lymphocyte ratio (PLR). These indices are widely reported as markers of cardiovascular risk, but it is unclear whether they provide comparable information or whether SII, which is derived from platelet count and NLR, adds anything beyond NLR. We analyzed two independent samples from the National Health and Nutrition Examination Survey (NHANES): 15,141 adults from 2005-2010 (development cohort) and 15,392 adults from 2011-2016 (validation cohort), each with complete blood count data and linked mortality follow-up. Survey-weighted Cox models, restricted cubic splines, multiple imputation, and model comparison were used, and the development model was applied to the validation cohort. During a median follow-up of 11.4 years, 795 cardiovascular deaths occurred in the development cohort; the validation cohort recorded 364 cardiovascular deaths over 5.75 years. In the development cohort, hazard ratios per 1-SD increase were 1.32 (95% CI 1.21-1.44) for NLR, 1.21 (1.12-1.32) for SII, and 1.13 (1.04-1.22) for PLR. When NLR and SII were entered together, only NLR remained associated (HR 1.49, 1.23-1.81). The validation cohort showed the same pattern: NLR was strongest (HR 1.35, 1.22-1.49), and neither SII nor PLR remained significant after mutual adjustment. The frozen model showed good discrimination (C-index 0.8991 to 0.9023) and calibration (slope 0.96). Adding NLR produced only a small improvement in discrimination, and reclassification results were inconsistent between cohorts.

**Author Summary:** Blood tests are commonly used to estimate inflammation, and several ratios calculated from a complete blood count, such as the neutrophil-to-lymphocyte ratio (NLR), systemic immune-inflammation index (SII), and platelet-to-lymphocyte ratio (PLR), have been proposed as markers of cardiovascular risk. Because SII is calculated from platelet count and NLR, it is important to know whether SII provides information beyond NLR. We studied this question in more than 30,000 US adults from two independent national surveys, with mortality follow-up through 2019. NLR was the strongest and most consistent marker in both samples. SII and PLR were associated with cardiovascular death when examined alone, but neither remained associated after NLR was taken into account. A model developed in the earlier survey also performed well in the later survey, with good discrimination and calibration. However, adding NLR to a model that already included established risk factors produced only a small improvement in prediction. These results suggest that SII should not be automatically preferred over NLR.

## Introduction

Cardiovascular disease remains a leading cause of death worldwide ^[1]^. Atherosclerosis is not simply a disorder of lipid storage; inflammation is involved from the earliest stages of endothelial injury through plaque rupture and thrombosis ^[2-4]^. Blood cells and platelets are easy to measure in routine practice, which has made inflammation-based ratios attractive as potential markers of cardiovascular risk ^[5]^.

NLR, PLR, and SII are calculated from the same complete blood count. NLR reflects the balance between neutrophils and lymphocytes ^[5]^; PLR combines platelet and lymphocyte counts ^[6,7]^; SII combines platelets, neutrophils, and lymphocytes and was first proposed for cancer prognosis ^[8]^. Each index has been linked to cardiovascular outcomes in particular populations, including people with hypertension ^[9]^, cancer survivors ^[10]^, patients with chronic kidney disease ^[11]^, patients with chronic obstructive pulmonary disease ^[12]^, and various cardiovascular conditions ^[13-16,17,18]^.

What remains unclear is whether these indices are interchangeable. Most studies have examined only one index and have assumed a linear relationship with outcome. Because SII is mathematically related to NLR, it is especially important to know whether SII adds information beyond NLR. We therefore compared the three indices in a large, nationally representative cohort with long-term mortality follow-up and then repeated the analyses in an independent sample from a later period.

## Materials and Methods

### Study design and population

We used data from NHANES 2005-2006, 2007-2008, and 2009-2010 ^[19]^. NHANES is a nationally representative, cross-sectional survey of the non-institutionalized US population conducted by the National Center for Health Statistics. Participants were linked to the public-use linked mortality file with follow-up through December 31, 2019 ^[20,21]^. Exposure and covariate data were collected during a single examination, and outcomes were ascertained by linking participants to subsequent mortality records. The present analysis should therefore be regarded as a cohort analysis of cross-sectional survey data with linked mortality follow-up, rather than a traditional prospective cohort study.

Adults aged 20 years or older with complete data on neutrophil count, lymphocyte count, and platelet count were eligible. Participants who were pregnant, who had missing components required for the calculation of NLR, PLR, or SII, who were not eligible for mortality follow-up, or who had no valid follow-up time were excluded. The analytic sample included 15,141 participants; the fully adjusted model included 13,591 participants with complete covariate data and 681 cardiovascular deaths.

### Exposure definitions

NLR was calculated as neutrophil count / lymphocyte count. PLR was calculated as platelet count / lymphocyte count. SII was calculated as platelet count × neutrophil count / lymphocyte count ^[8]^. All three indices were log-transformed and standardized; hazard ratios were expressed per 1-SD increase. Quartiles were derived using survey-weighted cut points, with the lowest quartile as the reference.

### Outcome ascertainment

The primary outcome was cardiovascular mortality, defined as death with an underlying cause of death coded as diseases of the heart or cerebrovascular diseases in the linked mortality file ^[20,21]^. All-cause mortality was a secondary outcome. Follow-up time was calculated from the examination date to death or December 31, 2019.

### Covariates

Covariates included age, sex, race/ethnicity, education, family income-to-poverty ratio, body mass index, smoking status, alcohol use, hypertension, diabetes, baseline cardiovascular disease, and estimated glomerular filtration rate (eGFR) calculated with the CKD-EPI 2021 equation without the race coefficient ^[22]^. Hypertension was defined as self-reported hypertension, systolic blood pressure ≥ 140 mmHg, or diastolic blood pressure ≥ 90 mmHg. Diabetes was defined as self-reported diabetes, HbA1c ≥ 6.5%, or fasting glucose ≥ 126 mg/dL. Baseline cardiovascular disease was defined as self-reported congestive heart failure, coronary heart disease, angina, myocardial infarction, or stroke.

### Statistical analysis

NHANES sampling weights, strata, and primary sampling units were incorporated using the survey package in R. For the three two-year cycles, combined weights were calculated according to the NHANES analytic guidelines, with the two-year examination weight divided by three ^[19]^. Survey-weighted Cox proportional hazards models were used. Three adjustment sets were constructed: model 1 (unadjusted), model 2 (age, sex, and race/ethnicity), and model 3 (model 2 plus BMI, education, income, hypertension, diabetes, smoking, alcohol, baseline cardiovascular disease, and eGFR). Restricted cubic splines with four knots were used to examine nonlinearity, and sensitivity analyses used three and five knots. Subgroup analyses were performed by age, sex, BMI, hypertension, diabetes, and baseline cardiovascular disease, with interaction tests. Sensitivity analyses excluded deaths within the first two years of follow-up and participants with baseline cardiovascular disease, and additional analyses used untransformed exposures. Missing covariates were handled with multiple imputation by chained equations (m = 5). Discrimination and reclassification were assessed using the C-index, continuous NRI, and IDI in exploratory analyses. Analyses were performed with R 4.4.1. Reporting followed STROBE ^[23]^.

### External validation cohort

An independent temporal validation cohort was constructed from NHANES 2011-2012, 2013-2014, and 2015-2016 using the same inclusion and exclusion criteria and the same variable definitions as the development cohort. The validation cohort included 15,392 participants; 364 cardiovascular deaths occurred during a median follow-up of 5.75 years. Complete covariate data were available for 13,642 participants (309 cardiovascular deaths). We performed two validation analyses: replication of the survey-weighted Cox models in the validation cohort, and application of the frozen development model to the validation cohort to assess discrimination (C-index) and 5-year calibration. Continuous NRI and IDI for the addition of NLR to the base model were also calculated in the validation cohort.

## Results

### Study population

A total of 15,141 adults were included; participant selection is shown in Figure 1. During follow-up, 795 cardiovascular deaths and 2,579 all-cause deaths occurred. The fully adjusted model included 13,591 participants and 681 cardiovascular deaths. The median follow-up was 11.4 years.

**Figure 1.**
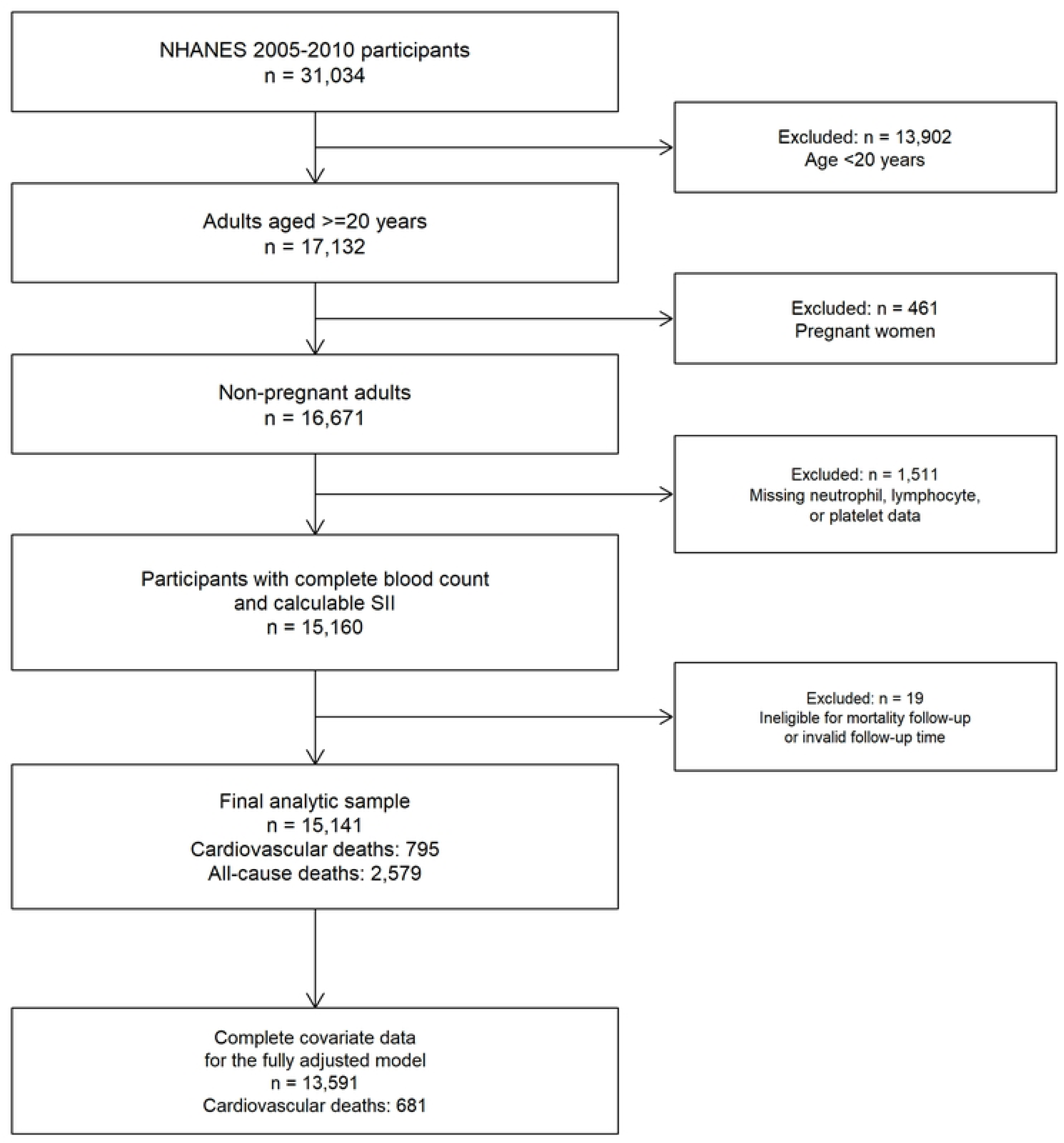
Flow diagram of participant selection.

**Figure 2.**
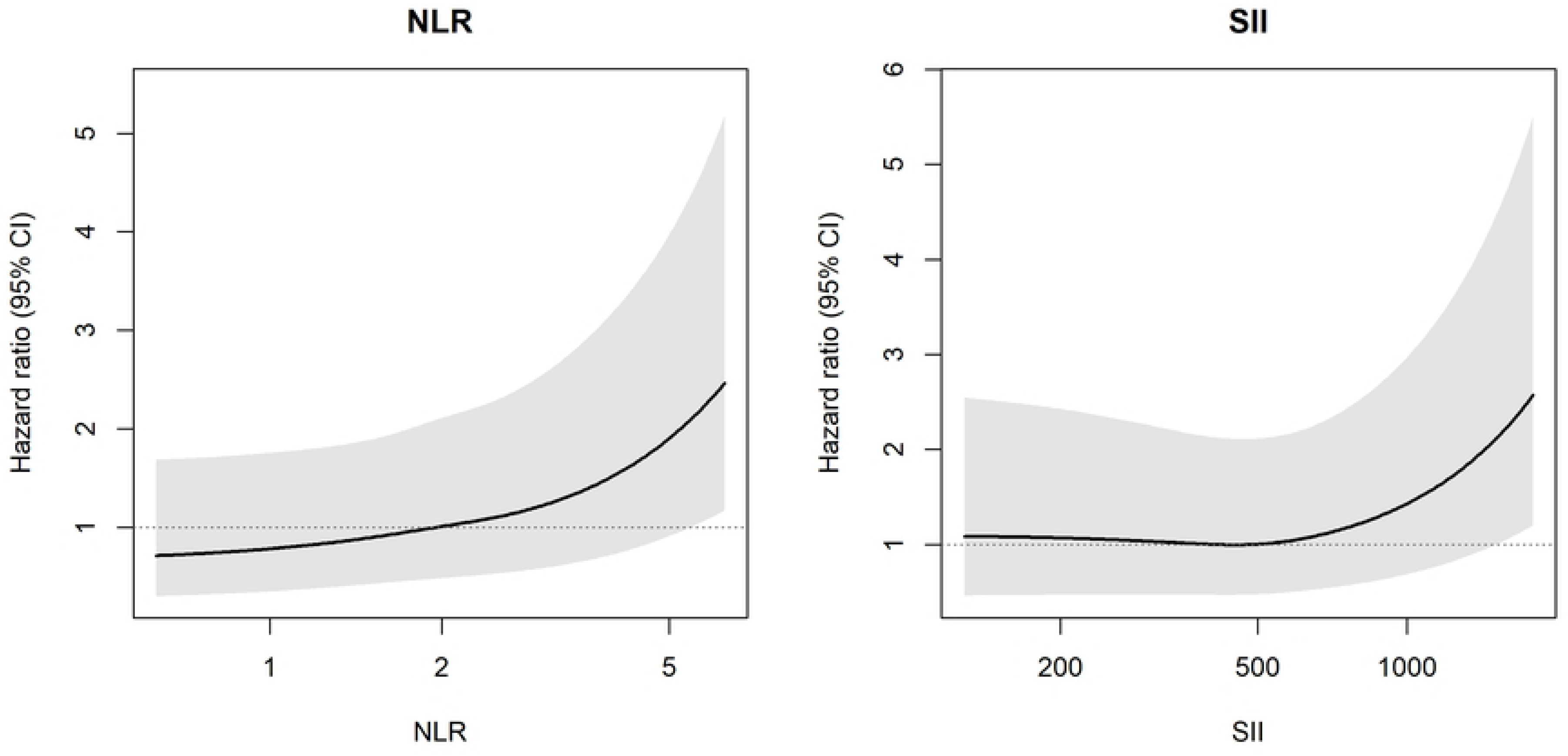
Restricted cubic spline curves for the associations of NLR and SII with cardiovascular mortality. Models were adjusted for age, sex, race/ethnicity, BMI, education, income-to-poverty ratio, hypertension, diabetes, smoking, alcohol use, baseline cardiovascular disease, and eGFR. The reference value was the median. The x-axis is shown on a logarithmic scale. Shaded areas represent 95% confidence intervals. The wide confidence intervals at the highest values indicate that estimates in the extreme upper tail are based on few participants and should be interpreted cautiously.

**Figure 3.**
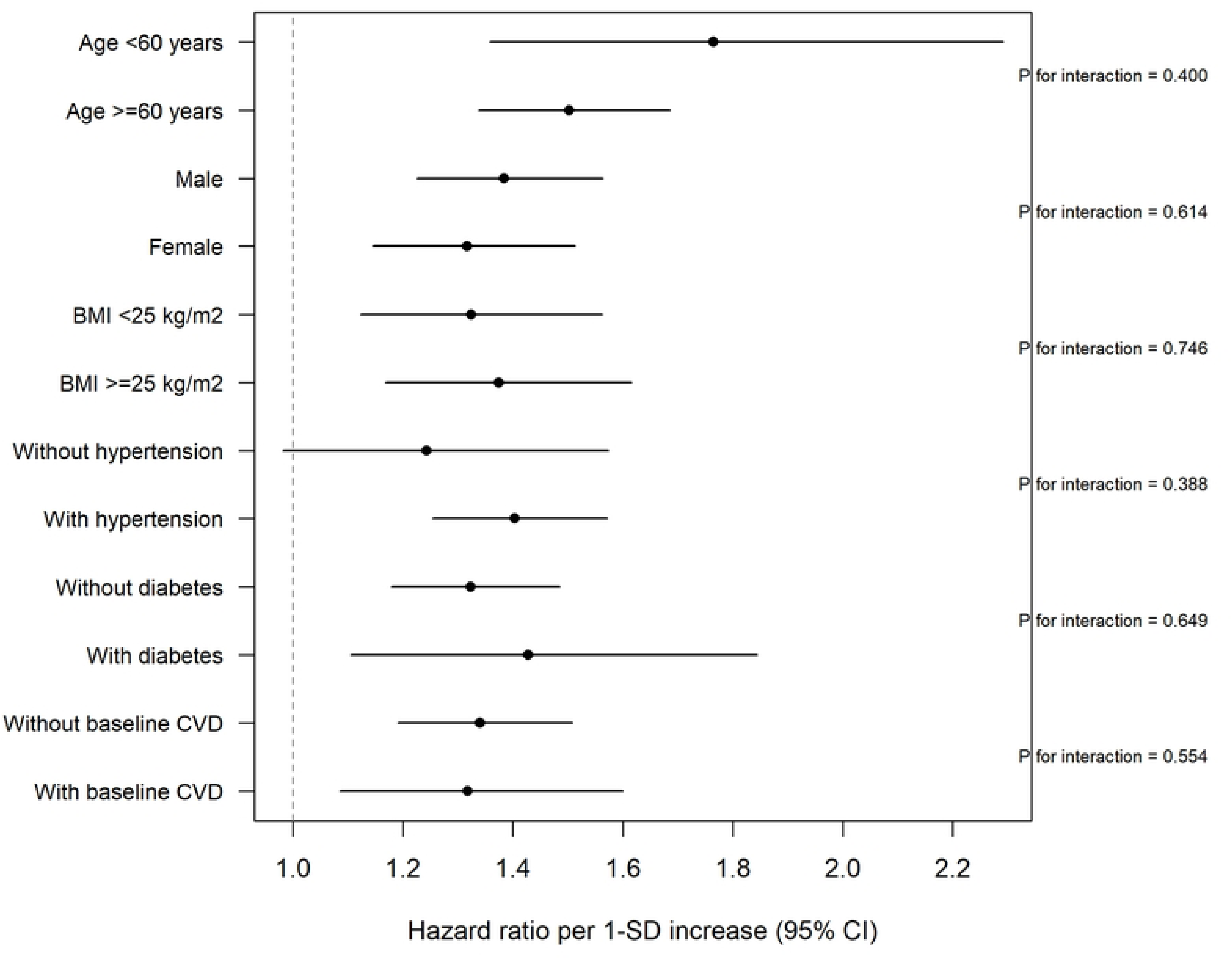
Subgroup analyses of the association between NLR and cardiovascular mortality. Hazard ratios are expressed per 1-SD increase in log NLR. Interaction P values are shown for each subgroup. No statistically significant interactions were observed; however, subgroup analyses may be underpowered.

Participants in the highest NLR quartile were older (mean age 50.4 vs 44.7 years), had a higher BMI (29.1 vs 28.0 kg/m^2^), lower eGFR (91.3 vs 97.2 mL/min/1.73 m^2^), and a higher prevalence of hypertension (43.1% vs 33.3%), diabetes (14.9% vs 10.5%), current smoking (24.4% vs 20.9%), and baseline cardiovascular disease (13.1% vs 6.3%) than those in the lowest quartile (Table 1). SII and PLR were also higher in the highest NLR quartile.

**Table 1.**
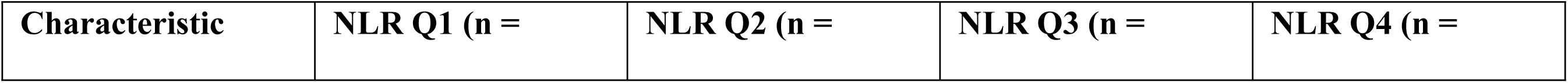
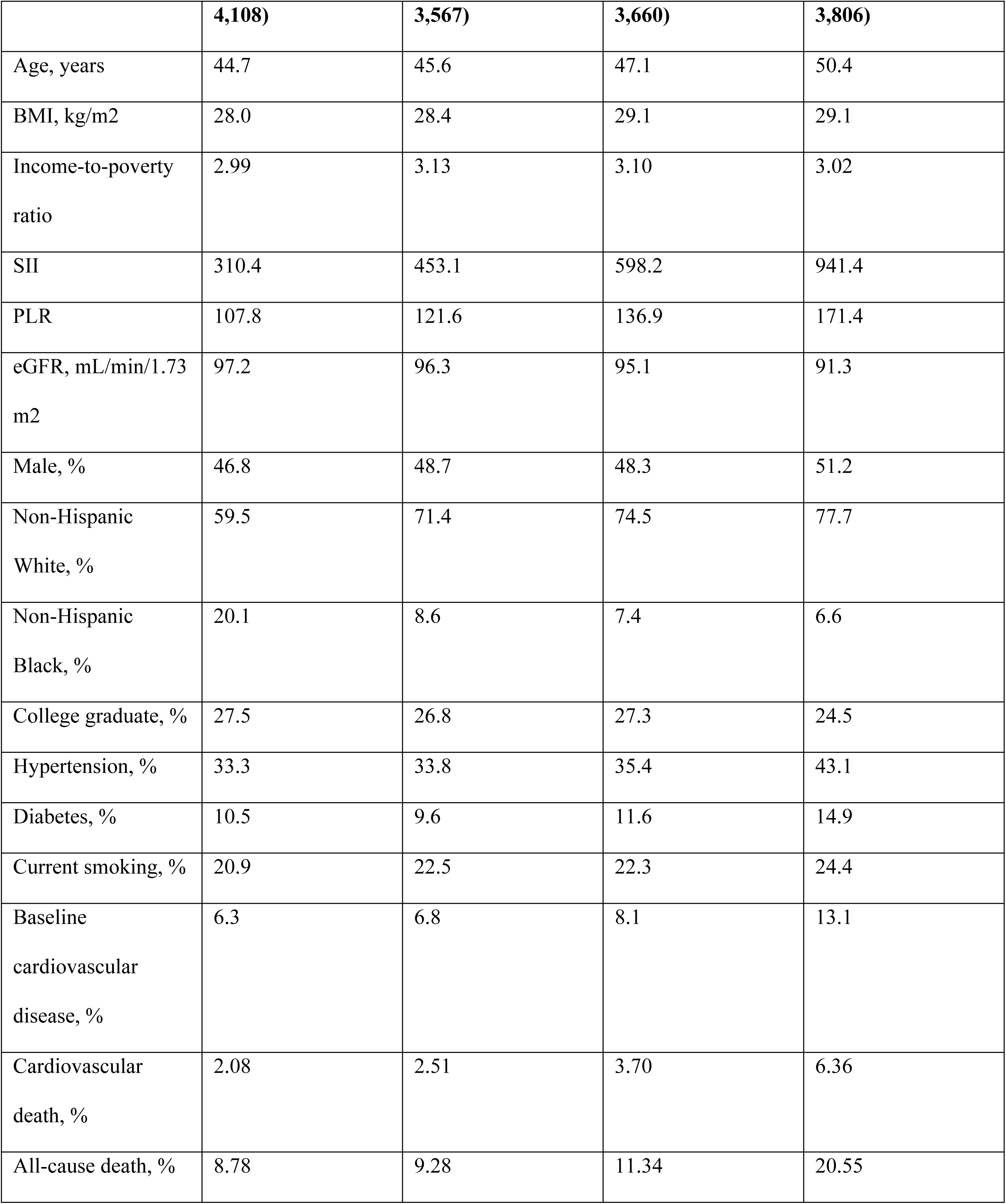
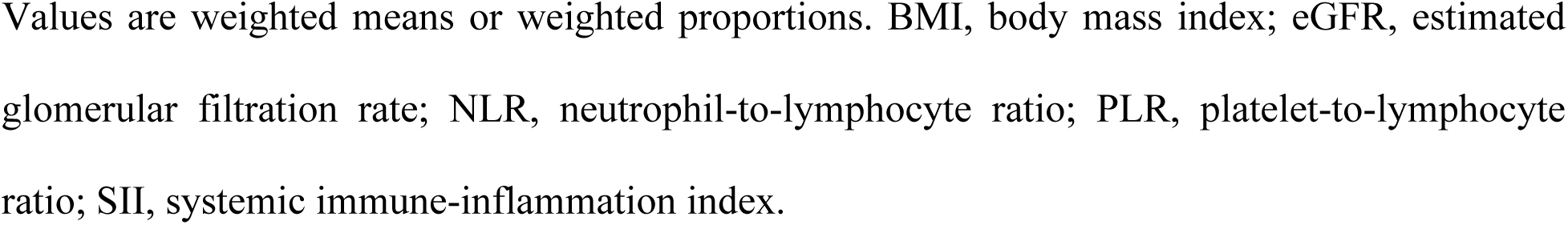
Baseline characteristics by NLR quartile. Values are weighted means or weighted proportions. BMI, body mass index; eGFR, estimated glomerular filtration rate; NLR, neutrophil-to-lymphocyte ratio; PLR, platelet-to-lymphocyte ratio; SII, systemic immune-inflammation index.

Cardiovascular deaths occurred in 133 of 4,108 participants in the lowest NLR quartile (weighted proportion 2.08%) and 338 of 3,806 participants in the highest quartile (6.36%). The corresponding weighted proportions for all-cause mortality were 8.78% and 20.55%. For SII, the weighted proportions of cardiovascular death were 3.00%, 2.86%, 3.64%, and 5.14% across increasing quartiles, and the weighted proportions of all-cause death were 10.77%, 10.42%, 11.60%, and 17.14%.

### Associations with cardiovascular mortality

In the fully adjusted models, each 1-SD increase in log NLR was associated with cardiovascular mortality (HR 1.32, 95% CI 1.16-1.51). The corresponding HRs were 1.22 (1.07-1.39) for SII and 1.13 (1.01-1.27) for PLR (Table 2).

**Table 2.** Associations of NLR, SII, and PLR with cardiovascular and all-cause mortality. Hazard ratios are expressed per 1-SD increase in the log-transformed index and are adjusted for age, sex, race/ethnicity, BMI, education, income-to-poverty ratio, hypertension, diabetes, smoking, alcohol use, baseline cardiovascular disease, and eGFR. CI, confidence interval; HR, hazard ratio; NLR, neutrophil-to-lymphocyte ratio; PLR, platelet-to-lymphocyte ratio; SII, systemic immune-inflammation index.

| Model | Index | Cardiovascular mortality, HR (95% CI) | All-cause mortality, HR (95% CI) |
| --- | --- | --- | --- |
| Single-index | NLR | 1.32 (1.16-1.51) | 1.19 (1.13-1.25) |
| Single-index | SII | 1.22 (1.07-1.39) | 1.12 (1.06-1.18) |
| Single-index | PLR | 1.13 (1.01-1.27) | 1.04 (0.98-1.10) |
| NLR + SII | NLR | 1.49 (1.23-1.81) | - |
| NLR + SII | SII | 0.87 (0.72-1.06) | - |
| NLR + PLR | NLR | 1.38 (1.21-1.58) | - |
| NLR + PLR | PLR | 0.94 (0.84-1.05) | - |
| All three | NLR | 1.49 (1.22-1.81) | - |
| All three | SII | 0.89 (0.71-1.11) | - |
| All three | PLR | 0.98 (0.86-1.12) | - |

**Table 3.**
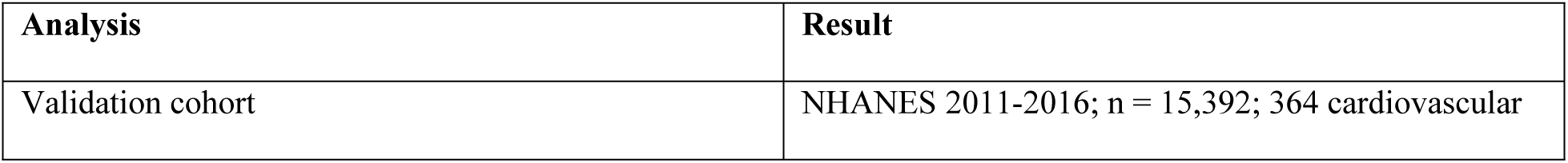
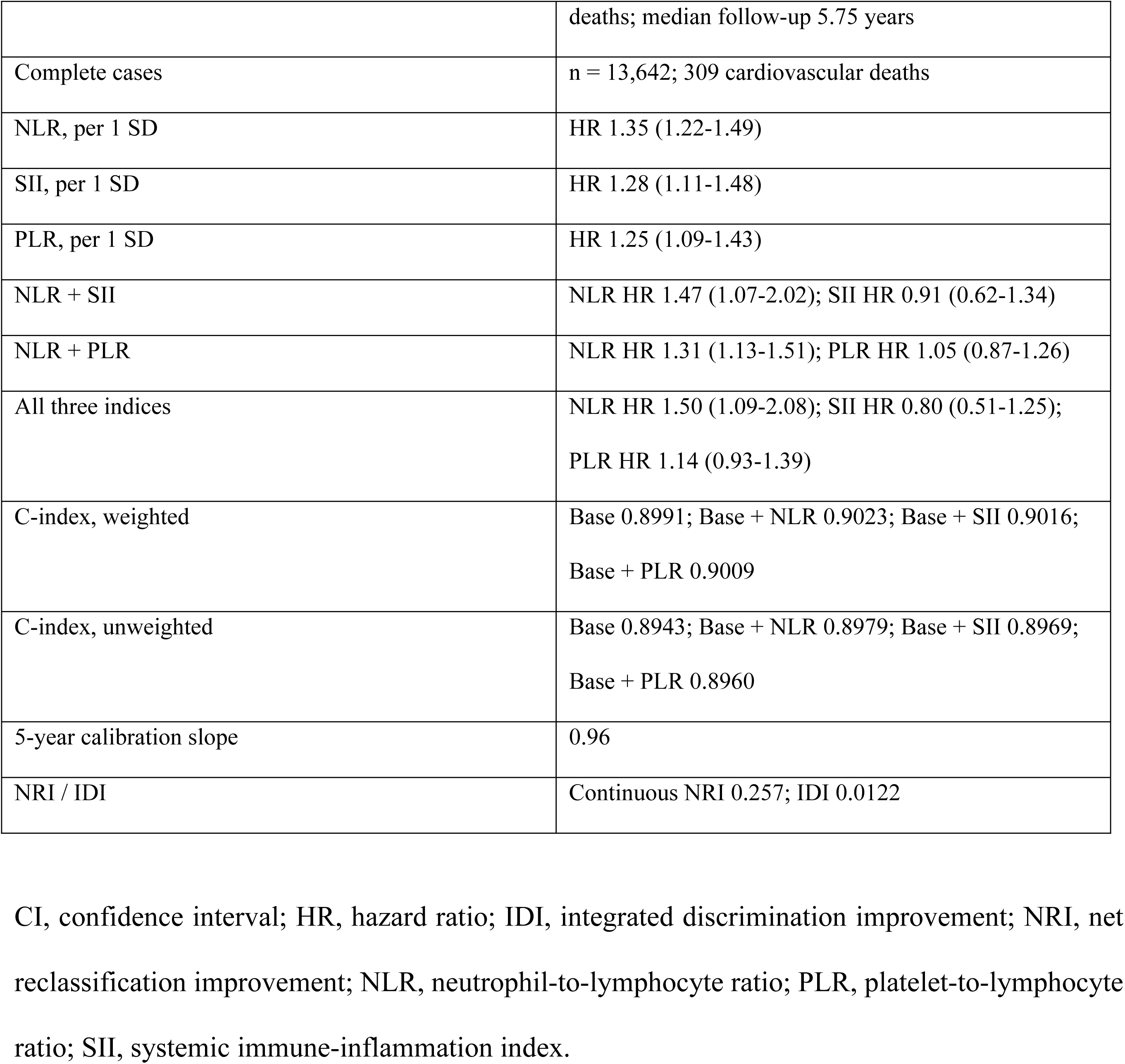
External validation of the inflammatory indices and the frozen model. CI, confidence interval; HR, hazard ratio; IDI, integrated discrimination improvement; NRI, net reclassification improvement; NLR, neutrophil-to-lymphocyte ratio; PLR, platelet-to-lymphocyte ratio; SII, systemic immune-inflammation index.

When NLR and SII were mutually adjusted, only NLR remained associated with cardiovascular mortality (HR 1.49, 1.23-1.81), whereas SII was not (HR 0.87, 0.72-1.06). Similarly, when NLR and PLR were combined, NLR remained associated (HR 1.38, 1.21-1.58) and PLR was not (HR 0.94, 0.84-1.05). In a model including all three indices, only NLR remained significant (HR 1.49, 1.22-1.81); SII (HR 0.89, 0.71-1.11) and PLR (HR 0.98, 0.86-1.12) were not associated.

### Nonlinearity and threshold effects

Restricted cubic spline analyses showed significant nonlinearity for both NLR (P = 3.6 × 10^-5^) and SII (P = 8.5 × 10^-6^). In fully adjusted models with the median as the reference, risk increased mainly in the highest percentile. At the 99th percentile, the HR was 2.47 (1.17-5.19) for NLR and 2.57 (1.20-5.52) for SII. In quartile analyses, only the highest quartile was significantly associated with cardiovascular mortality: NLR Q4 vs Q1 HR 1.69 (1.19-2.41); SII Q4 vs Q1 HR 1.39 (1.02-1.90).

### Subgroup analyses

No statistically significant interactions were observed across subgroups. HRs per 1-SD increase were 1.76 (1.36-2.29) in participants younger than 60 years and 1.50 (1.34-1.69) in those aged 60 years or older (interaction P = 0.40); 1.38 (1.23-1.56) in men and 1.32 (1.15-1.51) in women (interaction P = 0.61); 1.32 (1.12-1.56) in participants with BMI < 25 kg/m^2^ and 1.37 (1.17-1.62) in those with BMI ≥ 25 kg/m^2^ (interaction P = 0.75); 1.24 (0.98-1.57) in participants without hypertension and 1.40 (1.25-1.57) in those with hypertension (interaction P = 0.39); 1.32 (1.18-1.48) in participants without diabetes and 1.43 (1.11-1.84) in those with diabetes (interaction P = 0.65); and 1.34 (1.19-1.51) in participants without baseline cardiovascular disease and 1.32 (1.09-1.60) in those with baseline cardiovascular disease (interaction P = 0.55). These subgroup analyses may be underpowered, and the numerically stronger association in younger participants should be interpreted cautiously.

### Sensitivity and discrimination analyses

The NLR association was robust in sensitivity analyses: the HR was 1.30 (1.14-1.49) after excluding deaths within the first two years and 1.36 (1.20-1.53) after excluding participants with baseline cardiovascular disease. For all-cause mortality, the HR per 1-SD increase was 1.19 (1.13-1.25) for NLR, 1.12 (1.06-1.18) for SII, and 1.04 (0.98-1.10) for PLR.

In unweighted analyses, the C-index was 0.9008 for the base model, 0.9029 after adding NLR, 0.9021 after adding SII, and 0.9014 after adding PLR. Adding NLR and SII together yielded a C-index of 0.9030. Weighted analyses showed a similar pattern.

### Missing data and multiple imputation

Complete covariate data were available for 13,591 of 15,141 participants (681 of 795 cardiovascular deaths). Participants excluded from the complete-case analysis were older (mean age 48.3 vs 46.9 years), had a higher BMI (29.7 vs 28.6 kg/m^2^), a lower income-to-poverty ratio (2.72 vs 3.06), and higher proportions of cardiovascular death (5.75% vs 3.48%) and all-cause death (18.75% vs 11.92%). In multiple imputation analyses (m = 5), the results were consistent with the complete-case analysis. For cardiovascular mortality, the HRs per 1-SD increase were 1.32 (1.21-1.44) for NLR, 1.21 (1.12-1.32) for SII, and 1.13 (1.04-1.22) for PLR. For all-cause mortality, the corresponding HRs were 1.21 (1.15-1.26), 1.13 (1.08-1.18), and 1.05 (1.01-1.10), respectively.

### Model comparison and nonlinearity sensitivity

In unweighted models, adding NLR to the base model significantly improved model fit (likelihood ratio chi-square 32.29, df = 1, P = 1.3 × 10^-8^; AIC 10,835.74 vs 10,805.45), whereas adding SII on top of NLR did not (likelihood ratio chi-square 1.23, df = 1, P = 0.267; AIC 10,805.45 vs 10,806.22). Nonlinearity was robust to the number of knots. For NLR, the P values for nonlinearity were 6.9 × 10^-5^, 3.6 × 10^-5^, and 8.1 × 10^-5^ with 3, 4, and 5 knots, respectively; for SII, the corresponding P values were 1.6 × 10^-6^, 8.5 × 10^-6^, and 1.7 × 10^-5^.

### Joint models for all-cause mortality and raw-scale sensitivity

In joint models for all-cause mortality, NLR remained associated with mortality (NLR + SII: HR 1.34, 1.20-1.49; NLR + PLR: HR 1.27, 1.18-1.36; all three indices: HR 1.31, 1.18-1.46), whereas SII and PLR did not show positive associations (SII: HR 0.87-0.94; PLR: HR 0.91-0.93). These inverse associations are likely to reflect collinearity and were not interpreted as protective effects. Results were similar when the indices were analyzed on their original scale (NLR per SD: HR 1.24, 1.16-1.32; SII: HR 1.12, 1.09-1.16).

### Reclassification

In an exploratory analysis, adding NLR to the base model did not improve 10-year risk reclassification (continuous NRI -0.019; NRI -0.043 for events and 0.024 for non-events; IDI - 2.2 × 10^-5^; 663 cardiovascular deaths and 10,457 participants who remained event-free at 10 years).

### External validation

The validation cohort included 15,392 participants, with 364 cardiovascular deaths during a median follow-up of 5.75 years; 13,642 participants had complete covariate data (309 cardiovascular deaths). In the validation cohort, the fully adjusted HRs per 1-SD increase were 1.35 (1.22-1.49) for NLR, 1.28 (1.11-1.48) for SII, and 1.25 (1.09-1.43) for PLR. In joint models, only NLR remained associated (NLR + SII: NLR HR 1.47, 1.07-2.02; SII HR 0.91, 0.62-1.34; NLR + PLR: NLR HR 1.31, 1.13-1.51; PLR HR 1.05, 0.87-1.26; all three indices: NLR HR 1.50, 1.09-2.08; SII HR 0.80, 0.51-1.25; PLR HR 1.14, 0.93-1.39). When the frozen development model was applied to the validation cohort, the weighted C-index was 0.8991 for the base model and 0.9023 after adding NLR (unweighted: 0.8943 and 0.8979, respectively). The 5-year calibration slope was 0.96, and predicted risks were broadly consistent with observed risks across deciles, with slight underestimation in the highest decile (predicted 11.0% vs observed 16.2%). In the validation cohort, adding NLR produced a continuous NRI of 0.257 and an IDI of 0.0122, whereas these estimates were close to null in the development cohort.

## Discussion

We compared three inflammation-based indices in two separate NHANES samples and found that NLR was consistently the strongest marker of cardiovascular mortality. SII was associated with cardiovascular death when analyzed alone, but this association did not persist after adjustment for NLR. PLR behaved similarly: it was weakly associated in single-index models and lost significance when NLR was included. The validation cohort reproduced this pattern, which suggests that the relative performance of these indices is stable across different NHANES periods.

The overlap between SII and NLR is not surprising. SII is calculated as platelet count x NLR, so the two indices share the neutrophil and lymphocyte components. In our data, the weighted correlation between log SII and log NLR was 0.861. When both were included in the same model, the coefficient for SII became non-significant and, in some models, changed direction, a pattern typical of collinearity. For this reason, the joint model should be interpreted as showing that the two indices contain largely overlapping information, not as proof that SII has no prognostic value. SII may still be useful in other clinical settings, such as oncology or critical care, where it was originally developed and where the balance between inflammation and immune response may differ.

The nonlinear pattern also deserves attention. Both NLR and SII showed a threshold-like relationship with cardiovascular mortality, with risk increasing mainly in the upper tail of the distribution. In the development cohort, only the highest quartile was associated with a significant increase in risk. This pattern is consistent with the idea that mild elevations in these ratios may reflect normal variation, whereas marked elevations may indicate a sustained inflammatory state that promotes cardiovascular events. It also suggests that simple linear models may underestimate the effect at the high end and overstate it at the low end.

The clinical implications need to be stated carefully. NLR was statistically associated with cardiovascular mortality, and the association remained in the validation cohort. However, adding NLR to a model that already contained age, sex, race/ethnicity, socioeconomic factors, BMI, hypertension, diabetes, smoking, alcohol use, baseline cardiovascular disease, and eGFR produced only a small improvement in discrimination. In the development cohort, the addition of NLR did not improve reclassification, and the results in the validation cohort were inconsistent. Taken together, these findings argue against using NLR, SII, or PLR as stand-alone risk prediction tools. They may still be useful as inexpensive markers that can be obtained from routine blood tests, but their role in clinical decision-making remains to be defined.

Our study has several limitations. NHANES is a cross-sectional survey, and the present analysis is a cohort study of survey data linked to mortality records rather than a traditional prospective cohort. Residual confounding is possible; acute infection, chronic inflammatory conditions, and medications such as statins or corticosteroids were not fully captured. Blood counts were measured at a single time point, so we could not assess changes over time. Complete-case and multiple-imputation analyses gave consistent results, but selection bias cannot be entirely excluded if data were missing not at random. The validation cohort came from the same national survey system, and validation in a different population or health system would provide stronger evidence. Finally, the reclassification analyses were exploratory, and the C-index, NRI, and IDI should not be overinterpreted.

## Conclusions

NLR was the most consistent of the three indices in both the development and validation cohorts. SII did not add meaningful information beyond NLR, and PLR was weaker still. All three indices were associated with cardiovascular mortality to some degree, but their incremental value for risk prediction was small. NLR deserves further study as a simple, widely available marker, but it is not yet ready to replace established risk assessment tools.

## Data Availability

the present study used publicly available, de-identified NHANES data.

## Supporting Information

S1 Table. Quartile analyses, nonlinearity tests, sensitivity analyses, and discrimination of models with inflammatory indices.

S2 Table. Baseline characteristics by SII and PLR quartiles.

S3 Table. Model comparison, RCS knot sensitivity, multiple imputation, and reclassification analyses.

The STROBE checklist is provided as S1 Checklist.

## Data Availability

All data used in this study are publicly available from the National Center for Health Statistics (https://www.cdc.gov/nchs/nhanes/). The linked mortality files are available at https://www.cdc.gov/nchs/data-linkage/mortality-public.htm. The analysis code is provided as S1 File.

## Author Contributions

Conceptualization: Pengfei Guo, Haoyu Hu, Sumin Yang. Data curation: Pengfei Guo, Wenlong Yan. Formal analysis: Pengfei Guo. Investigation: Yuan An. Methodology: Pengfei Guo, Haoyu Hu. Project administration: Sumin Yang. Supervision: Sumin Yang. Visualization: Wenlong Yan. Writing - original draft: Pengfei Guo. Writing - review & editing: Pengfei Guo, Wenlong Yan, Haoyu Hu, Yuan An, Yalin Liu, Sumin Yang.

## Acknowledgments

The authors thank the participants and staff of the National Health and Nutrition Examination Survey for their contributions to the survey.

## Ethics Statement

NHANES is a publicly available, de-identified dataset. The present study did not require additional institutional review board approval. All procedures were conducted in accordance with the ethical standards of the institutional and national research committees and with the 1964 Helsinki Declaration and its later amendments.

## References

1. Roth GA, Mensah GA, Johnson CO, et al. Global Burden of Cardiovascular Diseases and Risk Factors, 1990-2019: Update From the GBD 2019 Study. J Am Coll Cardiol. (2020) 76(25):2982–3021.

2. Ajoolabady A, Pratico D, Lin L, et al. Inflammation in atherosclerosis: pathophysiology and mechanisms. Cell Death Dis. (2024) 15(11):817.

3. Libby P. The changing landscape of atherosclerosis. Nature. (2021) 592(7855):524–533.

4. Wolf D, Ley K. Immunity and Inflammation in Atherosclerosis. Circ Res. (2019) 124(2):315–327.

5. Bhat T, Teli S, Rijal J, et al. Neutrophil to lymphocyte ratio and cardiovascular diseases: a review. Expert Rev Cardiovasc Ther. (2013) 11(1):55–59.

6. Kurtul A, Ornek E. Platelet to Lymphocyte Ratio in Cardiovascular Diseases: A Systematic Review. Angiology. (2019) 70(9):802–818.

7. Serra R, Ielapi N, Licastro N, et al. Neutrophil-to-lymphocyte Ratio and Platelet-to-lymphocyte Ratio as Biomarkers for Cardiovascular Surgery Procedures: A Literature Review. Rev Recent Clin Trials. (2021) 16(2):173–179.

8. Hu B, Yang XR, Xu Y, et al. Systemic immune-inflammation index predicts prognosis of patients after curative resection for hepatocellular carcinoma. Clin Cancer Res. (2014) 20(23):6212–6222.

9. Zhang X, Wei R, Wang X, et al. The neutrophil-to-lymphocyte ratio is associated with all-cause and cardiovascular mortality among individuals with hypertension. Cardiovasc Diabetol. (2024) 23(1):117.

10. He Y, Liu X, Wang M, et al. Neutrophil-to-lymphocyte ratio as a predictor of cardiovascular mortality in cancer survivors. Sci Rep. (2024) 14(1):20980.

11. Zhao WM, Tao SM, Liu GL. Neutrophil-to-lymphocyte ratio in relation to the risk of all-cause mortality and cardiovascular events in patients with chronic kidney disease: a systematic review and meta-analysis. Ren Fail. (2020) 42(1):1059–1066.

12. Lan CC, Su WL, Yang MC, et al. Predictive role of neutrophil-percentage-to-albumin, neutrophil-to-lymphocyte and eosinophil-to-lymphocyte ratios for mortality in patients with COPD: Evidence from NHANES 2011-2018. Respirology. (2023) 28(12):1136–1146.

13. Xia Y, Xia C, Wu L, et al. Systemic Immune Inflammation Index (SII), System Inflammation Response Index (SIRI) and Risk of All-Cause Mortality and Cardiovascular Mortality: A 20-Year Follow-Up Cohort Study of 42,875 US Adults. J Clin Med. (2023) 12(3):1128.

14. Li W, Wang X, Diao H, et al. Systemic immune inflammation index with all-cause and cause-specific mortality: a meta-analysis. Inflamm Res. (2024) 73(12):2199–2216.

15. Lu Y, Wei M, Zhang K, et al. Systemic immune-inflammation index as a predictor of statin residual cardiovascular risk compared with C-reactive protein. Int J Cardiol. (2025) 438:133529.

16. Xu B, Wu Q, La R, et al. Is systemic inflammation a missing link between cardiometabolic index with mortality? Evidence from a large population-based study. Cardiovasc Diabetol. (2024) 23(1):212.

17. Hu X, Cheng S, Du H, et al. Association of platelet-to-lymphocyte ratio with 1-year all-cause mortality in ICU patients with heart failure. Sci Rep. (2024) 14(1):32016.

18. Tek M, Çetin MS, Diker E, et al. Platelet to lymphocyte ratio predicts all-cause mortality in patients with carotid arterial disease. Rom J Intern Med. (2019) 57(2):159–165.

19. Centers for Disease Control and Prevention, National Center for Health Statistics. National Health and Nutrition Examination Survey. Available at: https://www.cdc.gov/nchs/nhanes/ (accessed September 12, 2026).

20. Yankey BA, Rothenberg R, Strasser S, et al. Effect of marijuana use on cardiovascular and cerebrovascular mortality: A study using the National Health and Nutrition Examination Survey linked mortality file. Eur J Prev Cardiol. (2017) 24(17):1833–1840.

21. National Center for Health Statistics. Public-use linked mortality files. Available at: https://www.cdc.gov/nchs/data-linkage/mortality-public.htm (accessed September 12, 2026).

22. Inker LA, Eneanya ND, Coresh J, et al. New Creatinine- and Cystatin C-Based Equations to Estimate GFR without Race. N Engl J Med. (2021) 385(19):1737–1749.

23. von Elm E, Altman DG, Egger M, et al. The Strengthening the Reporting of Observational Studies in Epidemiology (STROBE) statement: guidelines for reporting observational studies. J Clin Epidemiol. (2008) 61(4):344–349.

